# Artificial Intelligence-Enabled Cardiac Function Estimation from Phone Videos of Echocardiograms

**DOI:** 10.64898/2026.06.19.26356088

**Authors:** Dhawal Modi, Jay Kim, Alexander Ye, Sahir Eusuff, Hirotaka Ieki, Andrew P. Ambrosy, Alan C. Kwan, Bryan He, James Zou, Susan Cheng, Euan Ashley, David Ouyang

## Abstract

**Importance:** Mobile phone-recorded echocardiogram videos are commonly used in point-of-care, telemedicine, and resource-limited workflows, but artificial intelligence models for left ventricular ejection fraction (LVEF) estimation have primarily been evaluated on native Digital Imaging and Communications in Medicine (DICOM) videos.

**Objective:** To evaluate whether previously described artificial intelligence models for LVEF estimation retain performance when applied to mobile phone-recorded echocardiographic videos.

**Design:** Multicenter model validation study comparing model-estimated LVEF with clinician-reported LVEF.

**Setting:** Three medical centers: Kaiser Permanente Northern California, Beth Israel Deaconess Medical Center through MIMIC-IV-ECHO, and Cedars-Sinai Medical Center.

**Participants:** Source studies with clinician-reported LVEF and apical 4-chamber or apical 2-chamber views, yielding 6209 phone-recorded videos from 2648 studies and 2611 patients.

**Exposures:** Mobile phone recording of native echocardiographic videos and fine-tuning of pretrained models using mobile phone-recorded videos from the Kaiser Permanente Northern California training cohort.

**Main Outcomes and Measures:** Mean absolute error in ejection fraction percentage points, R² for continuous estimation, and area under the receiver operating characteristic curve for identifying ejection fraction greater than 50%.

**Results:** The study included 6209 mobile phone-recorded echocardiographic videos from 2648 studies and 2611 patients; the weighted mean age was 68.4 years, and 1031 patients were male (39.5%). Without phone-video fine-tuning, the primary model achieved a mean absolute error of 7.00 percentage points, coefficient of determination of 0.49, and area under the receiver operating characteristic curve of 0.91 on phone-recorded videos; corresponding native DICOM performance was 6.08 percentage points, 0.60, and 0.93, respectively. On the 2396-video fine-tuning evaluation cohort, fine-tuning improved primary model performance to a mean absolute error of 6.96 percentage points, coefficient of determination of 0.61, and area under the receiver operating characteristic curve of 0.93. Fine-tuning the public EchoNet-Dynamic model improved performance from 9.36 percentage points, 0.37, and 0.84 to 7.86 percentage points, 0.50, and 0.89, respectively. Progressive central zoom preprocessing degraded model performance.

**Conclusions and Relevance:** These findings suggest that artificial intelligence–assisted left ventricular ejection fraction estimation from mobile phone-recorded echocardiograms may be feasible when native image export is unavailable, although prospective evaluation is needed before clinical deployment.

**Key Points:** **Question:** Can artificial intelligence models developed for native echocardiographic video formats estimate left ventricular ejection fraction from mobile phone-recorded echocardiogram videos?

**Findings:** In this multicenter retrospective model-validation study of 6209 phone-recorded echocardiographic videos from 2648 studies, the primary pretrained model achieved a mean absolute error of 7.00 percentage points and area under the curve of 0.91 for identifying ejection fraction greater than 50%; phone-video fine-tuning improved performance and digital zoom degraded performance.

**Meaning:** Artificial intelligence-assisted ejection fraction estimation from phone-recorded echocardiograms may support point-of-care and telemedicine workflows when native image export is unavailable.

## Introduction

Left ventricular ejection fraction (LVEF) is one of the most widely used measures of cardiac function and plays a critical role in the diagnosis, risk stratification, and treatment guidance for heart failure and other cardiovascular diseases across all demographics.^1–3^ In routine practice, echocardiography (especially transthoracic echocardiography) is the most common imaging modality used to assess LVEF because of its non-invasive quality, portability, and low risk of ionization.^5–7^ However, LVEF interpretation can vary between readers and maybe limited by image quality and availability of trained personnel, particularly in point-of-care or resource constrained settings.

Recent deep-learning approaches have shown that LVEF can be estimated directly from echocardiographic videos with accurate performance when compared to clinician assessment.^9,10^ Prior evaluations have been performed on native echocardiographic DICOM videos stored on the computer, however many clinicians do not have direct access to these times. In many clinical workflows, especially point-of-care ultrasound, telemedicine^12^ and low-resource settings, echocardiogram studies may instead be reviewed or shared as mobile phone-recorded videos of computer screens rather than exported as native DICOM files. These recordings can introduce compression artifacts, glare, motion, altered contrast, reframing and digital zoom, raising uncertainty about whether models trained on standard echocardiographic training data inputs remain reliable in this practical acquisition setting.

In this study, we evaluated existing deep-learning models for automated LVEF estimation on a dataset of 6,209 unique mobile phone-recorded echocardiogram videos from 2,648 studies across three geographically distinct medical centers. The overall study workflow is summarized in eFigure 1 in Supplement. We sought to determine whether a model trained for conventional echocardiogram DICOM videos can retain useful performance on mobile phone-recorded videos and identify any acquisition conditions under which performance degrades.

## Methods

### Dataset and Cohort

This study evaluated the performance of an automated LVEF estimation model on mobile phone-recorded echocardiographic videos. The source echocardiogram DICOMs were obtained from the MIMIC-IV-ECHO,^13^ KPNC and CSMC. Source echocardiograms were obtained from February 2025 at KPNC, 2014 - 2022 at CSMC, and 2017 – 2019 in MIMIC-IV-ECHO. Mobile phone recordings were generated from February 2026 to March 2026, and analyses were performed from March 2026 to April 2026. No longitudinal follow-up was performed. To simulate a practical mobile capture workflow, each source echocardiographic DICOM was displayed in MicroDicom Viewer and recorded using either an iPhone 11, iPhone 14, iPhone 16 Pro or Samsung Galaxy Ultra S25 mobile phone for the full duration of the video. Playback was performed at the recommended cine rate specified in the source DICOM metadata to preserve the temporal characteristics of the original echocardiogram as closely as possible.

We evaluated two previously published deep-learning models for automated LVEF estimation ^9,10^. For each phone video, videos were resized to 112 × 112 pixels and sampled as 32-frame clips with a temporal period of two frames, corresponding to approximately 15 frames/s for the 30 frames/s mobile phone recordings. A filtered manifest was used to identify source echocardiographic studies with clinician-assigned LVEF and to link each source DICOM to its corresponding mobile phone-recorded MP4 file by filename stem. After matching and deduplication, the final cohort comprised 6,209 unique videos corresponding to 2,648 unique studies. Clinician-assigned LVEF from the source echocardiogram was used as the reference ground truth for all analyses.

The cohort was also assigned view-specific labels by using an automated view-classification pipeline run on the original DICOMs. Only apical four chamber (A4C) and apical two chamber (A2C) videos were retained in the cohort as these views are the most relevant for LVEF estimation. This yielded a final inference dataset composed of mobile phone-recorded videos paired with clinician reference LVEF and categorized by apical views. Cohort derivation and the fine-tuning evaluation split are summarized in eFigure 2 in Supplement.

### Finetuning Strategy

In fine tuning experiments, we initialized the video-based LVEF regression model weights and fine-tuned it on mobile phone-recorded echocardiography videos from the KPNC cohort dataset paired with the clinical echocardiogram report LVEF. Splitting was performed at the study level to prevent leakage across train, validation, and test sets, using an 80:10:10 KPNC train/validation/internal-test split. The BIDMC and CSMC mobile phone video cohort was reserved as an external validation set and was not used for finetuning, model selection or hyperparameter tuning.

During training and validation, one randomly sampled clip per video was used per epoch, matching the checkpoint-selection strategy used previously. Pixel normalization statistics were computed from the Kaiser training split only and stored with the fine-tuned checkpoint. The model was optimized using mean squared error loss with stochastic gradient descent, momentum 0.9, weight decay 1 × 10^-4, and an initial learning rate of 1 × 10^-5 with stepwise decay. The checkpoint with the lowest validation mean squared error was selected for subsequent evaluation on the held-out KPNC test set, MIMIC and CSMC validation sets.

### Inferencing Pipeline

For each matched mobile phone-recorded video, the frame rate was obtained directly from the MP4 file and an integer temporal sampling period was calculated adaptively to approximate the temporal resolution expected by the training model. Videos were processed using the full frame without explicit cropping to the ultrasound region. During preprocessing, frames were centrally cropped and resized to 112 x 112 pixels to match the model input resolution. Pixel values were then normalized using the model’s precomputed channel-wise mean and standard deviation. Each mobile phone-recorded video was divided into non-overlapping fixed-length clips. A clip-level LVEF prediction was generated for each clip, and final clips shorter than the required clip length were excluded. The final video-level prediction was computed as the arithmetic mean of all clip-level predictions from that recording.

### Statistical analysis

Model performance was evaluated at the video level. MAE was calculated as the mean absolute difference between predicted and clinician-reported LVEF and is reported in LVEF percentage points. R² was used to quantify agreement between predicted and reference LVEF. AUC was calculated for classification of LVEF > 50%. Confidence intervals were calculated using nonparametric bootstrap resampling with 5000 iterations. Analyses were performed overall and stratified by site and acquisition subgroup.

## Results

We evaluated previously described AI-based LVEF estimation approaches^9,10^ on mobile phone-recorded echocardiographic videos and corresponding native DICOMs, including baseline and KPNC phone-video fine-tuned models. Performance was assessed on mobile phone-recorded videos, corresponding native DICOM, and across view, site, color/grayscale, and zoom subgroups. Table 1 summarizes the evaluation cohort, which included 6,209 mobile phone-recorded echocardiographic videos from 2,648 studies across KPNC, BIDMC and CSMC. Across the 2,611 patients represented in the cohort, the weighted mean age was 68.4 years, and 1,031 patients were male (39.5%). The cohort was racially and ethnically diverse, including 1,627 White patients (62.3%), 313 Black patients (12.0%), 267 Asian patients (10.2%), and 350 patients with Hispanic ethnicity (13.4%). Common comorbidities included hypertension in 1,545 patients (59.2%) and atrial fibrillation in 783 patients (30.0%). Site-level median LVEF ranged from 57% to 61%.

**Table 1.** Clinical characteristics of test sets. Values are presented as No., mean (SD), or median (IQR), unless otherwise indicated. Race and ethnicity were obtained from clinical records; Hispanic ethnicity is reported separately from race. Heart rate is reported in beats per minute. LVEF refers to clinician-reported left ventricular ejection fraction from the source echocardiogram. BIDMC, Beth Israel Deaconess Medical Center; BMI, body mass index; BSA, body surface area; CSMC, Cedars-Sinai Medical Center; KPNC, Kaiser Permanente Northern California; LVEF, left ventricular ejection fraction.

|  | **KPNC** | **BIDMC** | **CSMC** |
| --- | --- | --- | --- |
| **Patients, n** | 1184 | 198 | 1229 |
| **Studies, n** | 1184 | 223 | 1241 |
| **Videos, n** | 4247 | 715 | 1247 |
| **Age, years** | 68.1 (16.8) | 73.3 (12.5) | 68 (18.4) |
| **Males, n** | 630 | 103 | 298 |
| **Race, n** |  |  |  |
| **Asian** | 188 | 4 | 75 |
| **Black** | 101 | 34 | 178 |
| **Native American** | 5 | 0 | 4 |
| **Pacific Islander** | 5 | 0 | 3 |
| **White** | 664 | 120 | 843 |
| **Other/Unknown** | 221 | 40 | 126 |
| **Hispanic ethnicity, n** | 194 | 6 | 150 |
| **Height, mean (SD), m** | 1.68 (0.11) | 1.66 (0.10) | 1.69(0.13) |
| **Weight, mean (SD), kg** | 82.00 (23.40) | 78.88 (19.97) | 75.68 (20.87) |
| **BMI, mean (SD), kg/m<sup>2</sup>** | 28.6 (7.00) | 28.5 (6.3) | 26.25 (5.98) |
| **BSA, mean (SD), m<sup>2</sup>** | 1.90 (0.3) | 1.86 (0.25) | 1.87 (0.28) |
| **Hypertension, n** | 715 | 184 | 646 |
| **Atrial Fibrillation, n** | 372 | 81 | 330 |
| **Heart rate, 1/min** | 70 | 75 | 74 |
| **LVEF, mean (IQR), %** | 60 (53-64) | 57 (38-65) | 61 (52-66) |

### Model performance on phone-recorded videos and native DICOMs

The previously published model^9^ achieved a mean absolute error (MAE) of 7.00 percentage points (pp) [95% CI: 6.84 - 7.15], R^2^ of 0.49 [0.46 - 0.52] and AUC of 0.91 [0.90 - 0.92] for identifying LVEF > 50% (Figure 1). Site level performance is summarized in Table 2. On corresponding native DICOMs (original videos that phone recordings came from), the model achieved MAE of 6.08 pp [5.94 - 6.22], R^2^ 0.60 [0.57 - 0.62], and AUC 0.93 [0.92 - 0.93], compared with MAE 7.00 pp [6.84 - 7.15] and R² 0.49 [0.46 - 0.52] on phone-recorded videos.

**Figure 1.**
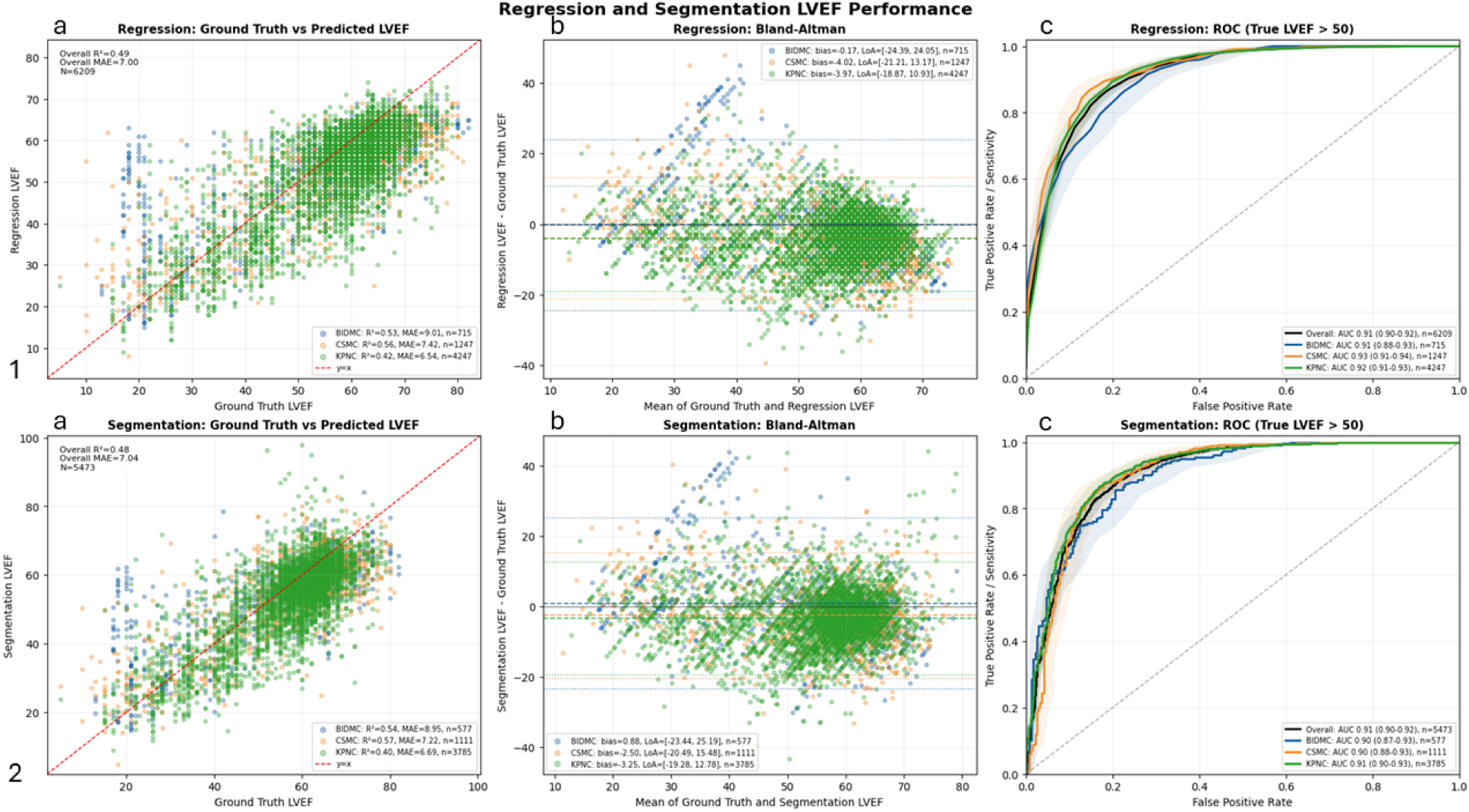
Row 1. Performance of automated LVEF estimation on mobile-phone-recorded echocardiographic videos. Subplot (A) Scatterplot comparing clinician-assigned ground-truth left ventricular ejection fraction (LVEF) with regression model-predicted LVEF for 6,209 unique mobile-phone-recorded echocardiographic videos in the primary no-zoom color condition for KPNC test cohort (Green), BIDMC test cohort (Blue) and CSMC test cohort (Yellow). The plot demonstrates moderate agreement between predicted and reference LVEF values, with greater dispersion at lower-to-mid LVEF values. Subplot (B) Bland-Altman plot showing agreement between model-predicted and clinician-assigned LVEF in the same cohort. The mean bias for KPNC test set (Green) was −3.97, indicating a modest overall tendency to underestimate LVEF. The 95% limits of agreement were −18.87 to 10.93. Similarly, mean bias for CSMC dataset (Yellow) was −4.02 and −0.17 for BIDMC dataset (Blue). Subplot (C) ROC curves for identifying echo videos with true LVEF > 50, shown overall and stratified by site. For KPNC test cohort (Green), BIDMC test cohort (Blue) and CSMC test cohort (Yellow). **Figure 1 Row 2. Performance of automated LVEF estimation using selected candidate frames from LV Segmentation model closest to corresponding regression model results on mobile-phone-recorded echocardiographic videos** Subplot (A) Scatterplot comparing clinician-assigned ground-truth left ventricular ejection fraction (LVEF) with LV Segmentation model-predicted LVEF for 5,473 unique mobile-phone-recorded echocardiographic videos in the primary no-zoom color condition for KPNC test cohort (Green), BIDMC test cohort (Blue) and CSMC test cohort (Yellow). Subplot (B) Bland-Altman plot showing agreement between model-predicted and clinician-assigned LVEF in the same cohort. The mean bias for KPNC test set (Green) was −3.25, similarly showing a pattern of underestimating LVEF. The 95% limits of agreement were −19.28 to 12.78. Similarly, mean bias for CSMC dataset (Yellow) was −2.50 and 0.88 for BIDMC dataset (Blue). Subplot (C) ROC curves for identifying studies with true LVEF > 50, shown overall and stratified by site for regression-guided segmentation derived LVEF estimation. For KPNC test cohort (Green), BIDMC test cohort (Blue) and CSMC test cohort (Yellow).

**Table 2.** Performance metrics of different LVEF models in different settings. Data are point estimates with 95% CIs in brackets. Performance was evaluated at the video level in the primary evaluation cohort. MAE is reported in LVEF percentage points. AUC was calculated for identifying clinician-reported LVEF greater than 50%. N indicates the number of videos. Clinician-reported LVEF from the source echocardiogram was used as the reference standard. AUC, area under the receiver operating characteristic curve; BIDMC, Beth Israel Deaconess Medical Center; CSMC, Cedars-Sinai Medical Center; DICOM, Digital Imaging and Communications in Medicine; KPNC, Kaiser Permanente Northern California; LVEF, left ventricular ejection fraction; MAE, mean absolute error; R², coefficient of determination.

| Models | Mobile Phone Videos |  |  |  |  |  |  |  |  |  |  |  |
| --- | --- | --- | --- | --- | --- | --- | --- | --- | --- | --- | --- | --- |
|  | Overall<br>(N = 6,209) |  |  | KPNC<br>(N = 4,247) |  |  | BIDMC<br>(N = 715) |  |  | CSMC<br>(N = 1,247) |  |  |
|  | R2 | MAE | AUC | R2 | MAE | AUC | R2 | MAE | AUC | R2 | MAE | AUC |
| He et al.<br>Nature<br>2023 | 0.49<br>[0.46 -<br>0.52] | 7.00<br>[6.84 -<br>7.15] | 0.91<br>[0.90 -<br>0.92] | 0.42<br>[0.38 -<br>0.46] | 6.54<br>[6.38 -<br>6.71] | 0.92<br>[0.91 -<br>0.93] | 0.53<br>[0.48 -<br>0.58] | 9.01<br>[8.39 -<br>9.62] | 0.91<br>[0.88 -<br>0.93] | 0.56<br>[0.51 -<br>0.61] | 7.42<br>[7.08 -<br>7.76] | 0.93<br>[0.91<br>0.94] |
| Ouyang<br>et al.<br>Nature<br>2020 | 0.33<br>[0.29 -<br>0.36] | 8.09<br>[7.92 -<br>8.26] | 0.86<br>[0.85 -<br>0.88] | 0.29<br>[0.24 -<br>0.34] | 7.30<br>[7.12 -<br>7.48] | 0.88<br>[0.87 -<br>0.89] | 0.33<br>[0.27 -<br>0.39] | 10.95<br>[10.28 -<br>11.67] | 0.83<br>[0.80 -<br>0.86] | 0.35<br>[0.27 -<br>0.42] | 9.15<br>[8.76 -<br>9.56] | 0.87<br>[0.84<br>0.89] |
|  | Native DICOM Videos |  |  |  |  |  |  |  |  |  |  |  |
| He et al.<br>Nature<br>2023 | 0.60<br>[0.57 -<br>0.62] | 6.08<br>[5.94 -<br>6.22] | 0.93<br>[0.92 -<br>0.93] | 0.53<br>[0.49 -<br>0.56] | 5.75<br>[5.59 -<br>5.91] | 0.92<br>[0.91 -<br>0.93] | 0.65<br>[0.59 -<br>0.70] | 7.61<br>[7.07 -<br>8.16] | 0.94<br>[0.92 -<br>0.95] | 0.66<br>[0.62 -<br>0.70] | 6.31<br>[6.01 -<br>6.63] | 0.94<br>[0.92<br>0.95] |
| Ouyang<br>et al.<br>Nature<br>2020 | 0.41<br>[0.37 -<br>0.44] | 7.40<br>[7.24 -<br>7.58] | 0.87<br>[0.86 -<br>0.89] | 0.35<br>[0.31 -<br>0.39] | 6.89<br>[6.71 -<br>7.07] | 0.88<br>[0.86 -<br>0.89] | 0.40<br>[0.32 -<br>0.47] | 9.91<br>[9.2 -<br>10.65] | 0.86<br>[0.83 -<br>0.89] | 0.49<br>[0.42 -<br>0.54] | 7.72<br>[7.33 -<br>8.11] | 0.88<br>[0.86<br>0.91] |

On the 2,396 video fine-tuning evaluation cohort, after fine-tuning on the KPNC training split, the model improved MAE to 6.96 pp [6.69 - 7.24] and R^2^ to 0.61 [0.59 - 0.64], and AUC of 0.93 [0.91 - 0.94] (Figure 2; Table 3). Site-level MAE improved at all sites; from 6.67 pp [6.14 - 7.24] to 5.67 pp [5.19 - 6.17] at KPNC, 9.01 pp [8.39 - 9.62] to 8.76 pp [8.15 - 9.37] at BIDMC, and 7.42 pp [7.08 −7.76] to 6.37 pp [6.05 - 6.70] at CSMC with similar improvements in R^2^ from 0.56 [0.46 - 0.64] to 0.67 [0.60 - 0.73] at KPNC, 0.53 [0.48 - 0.58] to 0.54 [0.48 - 0.59] at BIDMC and 0.56 [0.51 - 0.61] to 0.65 [0.61 - 0.68] at CSMC. Models trained on smaller cohorts (10k echocardiogram DICOM videos), also improved with fine-tuning. The original EchoNet-Dynamic model^10^ achieved MAE 8.09 pp [7.92 - 8.26], R^2^ 0.33 [0.29 - 0.36] and AUC 0.86 [0.85 - 0.88] on the phone-video cohort, and after fine-tuning, improved MAE from 9.36 pp [9.04 - 9.68] to 7.86 pp [7.54 - 8.18], R² from 0.37 [0.32 - 0.41] to 0.50 [0.46 - 0.53], and AUC from 0.84 [0.83 - 0.86] to 0.89 [0.88 - 0.91]. Site-level MAE improved from 7.36 pp [6.75 - 8.02] to 6.37 pp [5.78 - 6.99] at KPNC, 10.95 pp [10.25 - 11.67] to 9.77 pp [9.10 −10.48] at BIDMC, and 9.15 pp [8.73 - 9.56] to 7.28 pp [6.91 - 7.67] at CSMC (Figure 2; Table 3).

**Figure 2.**
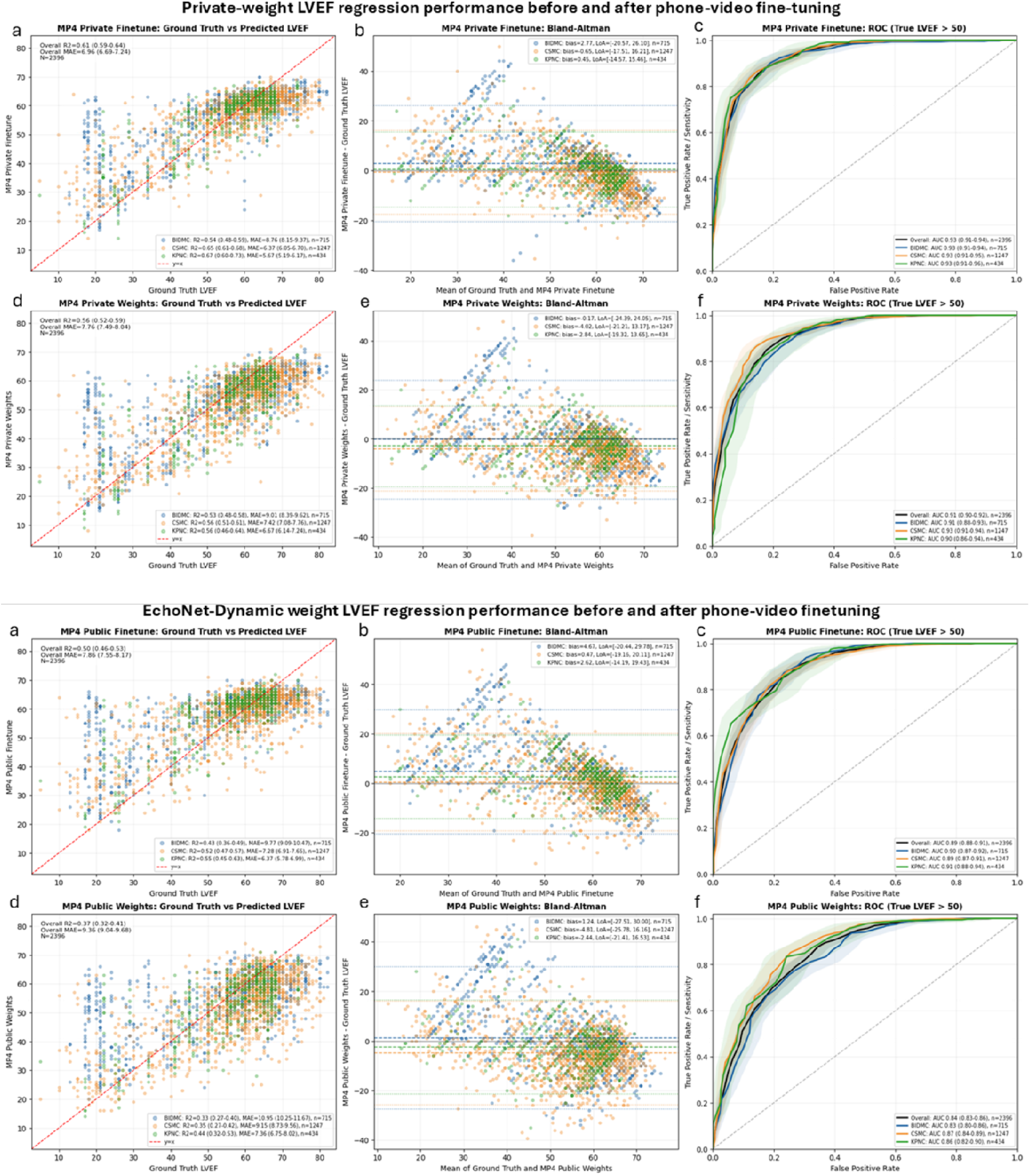
Change in LVEF Estimation Error Before and After Phone-Video Fine-Tuning. Mean absolute error (MAE) before and after phone-video fine-tuning is shown for the private-weight model and the public EchoNet-Dynamic model on the shared fine-tuning evaluation cohort. Panels show performance for private-weight phone-recorded videos (a), private-weight native Digital Imaging and Communications in Medicine (DICOM) videos (b), public-weight phone-recorded videos (c), and public-weight native DICOM videos (d). Points indicate MAE for the overall cohort and each validation site; connecting lines show the change after fine-tuning. MAE is reported in left ventricular ejection fraction percentage points. BIDMC indicates Beth Israel Deaconess Medical Center; CSMC, Cedars-Sinai Medical Center; KPNC, Kaiser Permanente Northern California.

**Table 3.**
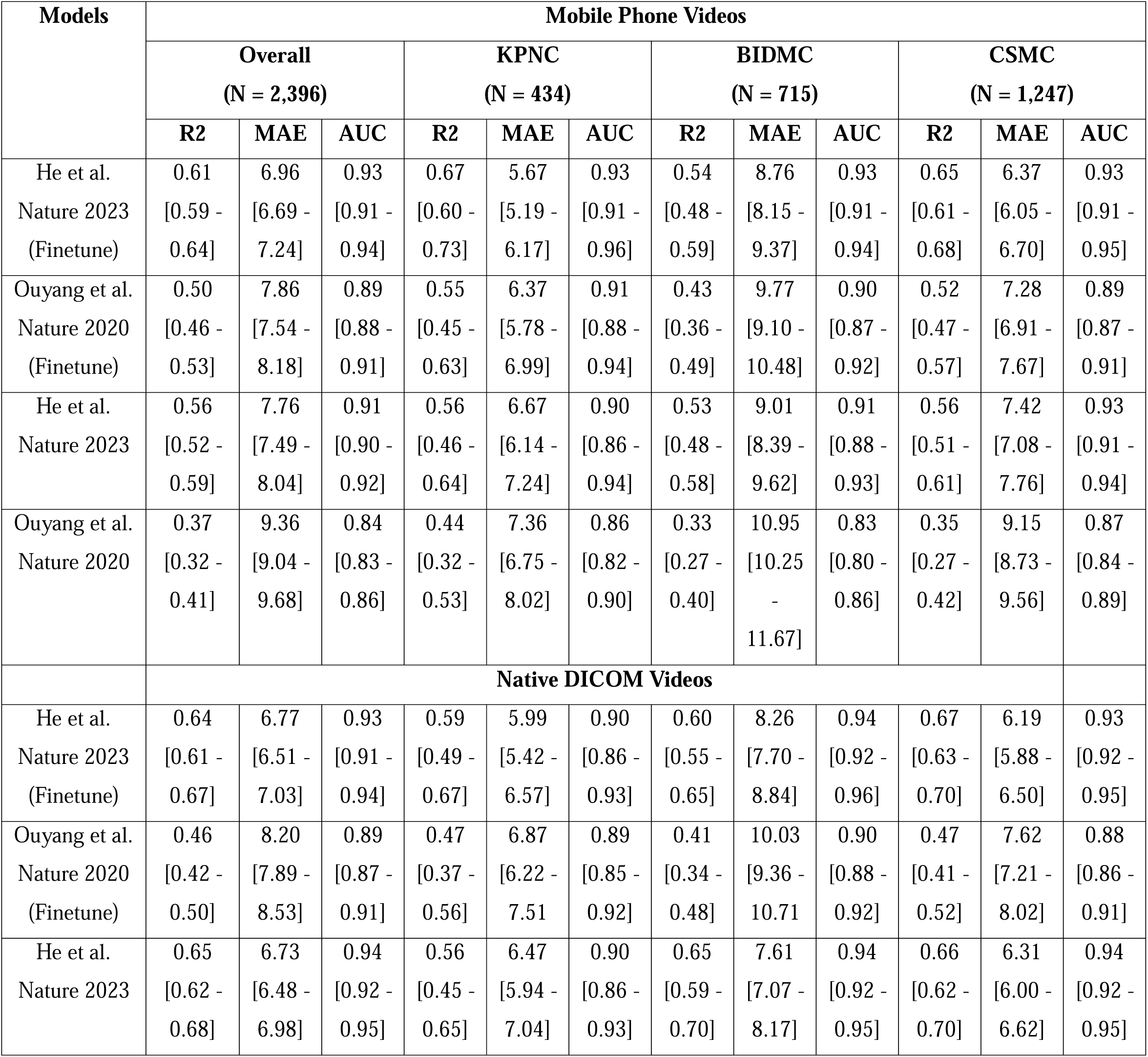
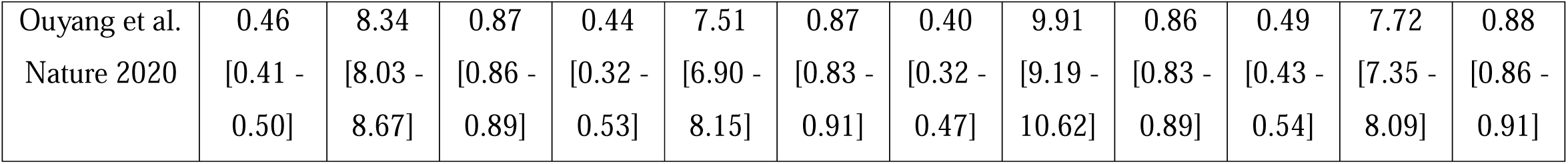
Performance metrics of Baseline and Finetuned LVEF model in different settings on test cohort. Data are point estimates with 95% CIs in brackets. The shared fine-tuning evaluation cohort included the held-out KPNC internal test set and the BIDMC and CSMC external validation cohorts. Fine-tuned models were trained using KPNC phone-recorded training and validation videos; BIDMC and CSMC videos were not used for fine-tuning, model selection, or hyperparameter tuning. MAE is reported in LVEF percentage points. AUC was calculated for identifying clinician-reported LVEF greater than 50%. N indicates the number of videos. AUC, area under the receiver operating characteristic curve; BIDMC, Beth Israel Deaconess Medical Center; CSMC, Cedars-Sinai Medical Center; DICOM, Digital Imaging and Communications in Medicine; KPNC, Kaiser Permanente Northern California; LVEF, left ventricular ejection fraction; MAE, mean absolute error; R^2^, coefficient of determination.

### Evaluating impact of acquisition parameters

In no-zoom color videos, regression model performance was similar between A4C and A2C views, with MAE / R² of 6.85 pp / 0.49 and 7.08 pp / 0.48, respectively. Color-to-grayscale conversion produced minimal change in overall performance, with MAE / R² of 7.00 pp / 0.49 for color videos and 7.07 pp / 0.48 for grayscale videos. Progressive zoom did not improve model performance. MAE increased from 7.00 pp at 0% zoom to 7.42 pp, 8.14 pp, and 9.01 pp at 10%, 20%, and 30% zoom, respectively. R² decreased from 0.49 to 0.43, 0.33, and 0.08 across the same zoom levels (eTable 1, eFigure 3 in Supplement).

## Discussion

Accurate and low-variability quantification of LVEF is critical for cardiac disease detection and management, as it allows for physicians to diagnose heart failure, determine its severity, and guide treatment.^1,2^ Although recent advancements in machine learning methods have demonstrated accurate performance for LVEF estimations,^9,10^ the practicality of implementing such models into clinical care must be considered. In low-resourced or rural settings, clinicians may share echocardiogram videos as mobile phone recordings due to limited infrastructure for direct transfer of radiologic imaging data. In this study, pretrained LVEF estimation models retained useful performance when applied to mobile phone-recorded echocardiographic videos supporting the feasibility of AI-assisted cardiac function assessment in point-of-care, telemedicine and resource limited settings.

The previously published model trained on DICOM images still had performance comparable to interobserver variability among clinicians for LVEF assessment,^8,9^ suggesting clinically acceptable accuracy despite the domain shift from native DICOM files to phone-recorded videos. The model also performed similarly across apical views and validation sites, suggesting that phone-recorded echocardiographic videos can preserve sufficient visual information for LVEF estimation. Importantly, threshold-based discrimination for LVEF >50% was stronger than continuous LVEF agreement, suggesting that phone-video inference may be most immediately useful for screening or triage of heart failure with reduced ejection fraction (HFrEF) rather than replacing formal quantitative echocardiographic interpretation.

Fine-tuning provided an additional practical strategy for adapting echocardiographic models to phone recorded videos. Both models trained on large and small corpuses of DICOMs improved after phone-video fine-tuning, supporting the value of domain specific adaptation when deploying models outside the native DICOM setting. Acquisition technique emerged as an important practical consideration. Color-to-grayscale conversion had minimal effect, whereas progressive central zoom/cropping degraded performance. These findings suggest that clear acquisition guidelines, particularly avoiding digital zoom and maintaining full-screen view, would be essential for clinical deployment of phone-based AI assessment.

Our study extends prior work demonstrating AI-based LVEF assessment from native echocardiographic data by addressing a critical gap in evaluating model robustness under real-world acquisition conditions^9,10^. Unlike previous studies that evaluated models exclusively on curated DICOM datasets, we assessed performance on videos subject to compression artifacts, variable screen display, and mobile phone capture. These are conditions representative of actual clinical practice in emergency, point-of-care, and resource-limited settings where formal DICOM export may be unavailable or impractical.

This study has limitations. First, mobile phone videos were generated by recording source echocardiographic DICOM videos displayed on a monitor, which may not capture the full variability of bedside or real-time clinical phone recording. Second, performance was evaluated at the video level, and multiple videos could originate from the same study or patient. Third, the cohort included only selected phone models, and three health systems, which may limit generalizability. Finally, this study evaluated retrospective model performance and did not assess prospective clinical workflow, clinician decision-making, or patient outcomes.

## Conclusions

This study assessed whether existing machine learning frameworks for LVEF estimation can generalize to echocardiograms acquired via alternative methods, specifically phone-recorded videos. While this approach is convenient for point-of-care use and vendor-agnostic deployment, image quality is constrained by factors such as compression, motion artifacts, and screen capture limitations, representing a significant domain shift. Previously published deep learning model for LVEF assessment maintains clinically useful performance when applied to mobile phone-recorded echocardiographic videos without retraining, achieving an accuracy similar to clinicians across three medical centers. Performance was robust to variations in phone model and video format but degraded substantially with digital zoom. These findings support the feasibility of mobile phone-based AI-assisted cardiac function assessment for point-of-care, emergency, and telemedicine applications where native DICOM access is unavailable.

## Declarations

### Data Availability

The KPNC and CSMC echocardiography datasets, derived mobile phone-recorded videos, linked clinical metadata, and study manifests cannot be publicly released because they contain protected health information and are subject to institutional data-use restrictions. The BIDMC source data were derived from MIMIC-IV-ECHO and are available to credentialed users through the MIMIC/PhysioNet data access process, subject to the applicable data use agreement.

### Code Availability

Code for this project is available at https://github.com/echonet/mobile-phone-videos

## Acknowledgements

This work has been supported by NIH NHLBI grants R00-HL157421, R01HL173487, and R01HL173526.

## Author Contributions

D.M. and D.O. conceived and designed the study. D.M. performed the primary analyses and drafted the manuscript. S.E, A.Y. and J.K. assisted with data curation, data processing and analysis. H.I., A.P.A., A.C.K., S.C., and E.A. provided clinical expertise and interpretation. B.H. provided methodological expertise and model resources. D.O. supervised the study. All authors reviewed and approved the final manuscript.

## Competing Interests

Dr. Ouyang has received research support from Alexion and AstraZeneca, as well as consulting fees or honoraria from Abbott, Johnson & Johnson, Dandelion Health, and AstraZeneca, and equity in InVision (all unrelated to the content of the present manuscript).

## Ethics Approval

This retrospective study was approved by the institutional review boards of KPNC and CSMC with waiver of informed consent because the study used retrospective echocardiographic data. The study followed applicable data-use agreements for MIMIC-IV-ECHO.

## Notes

### Competing Interest Statement

The authors have declared no competing interest.

