## Supplementary Data for "Artificial Intelligence-Enabled Cardiac Function Estimation from Phone Videos of Echocardiograms"

**Supplementary Data
Supplementary Tables**

**eTable 1 Performance comparison under different preprocessing zoom settings**

| **Cohort** | **Zoom Percentage** | | | | | | | |
| --- | --- | --- | --- | --- | --- | --- | --- | --- |
|  | **0%** | | **10%** | | **20%** | | **30%** | |
|  | **R^2^** | **MAE** | **R^2^** | **MAE** | **R^2^** | **MAE** | **R^2^** | **MAE** |
| Overall | 0.49 | 7.00 | 0.43 | 7.42 | 0.33 | 8.14 | 0.08 | 9.01 |
| KPNC | 0.42 | 6.54 | 0.36 | 6.97 | 0.27 | 7.55 | 0.11 | 7.98 |
| BIDMC | 0.53 | 9.01 | 0.45 | 9.72 | 0.33 | 10.95 | -0.09 | 13.89 |
| CSMC | 0.56 | 7.42 | 0.53 | 7.66 | 0.40 | 8.52 | 0.12 | 9.72 |

**Supplementary Figures**

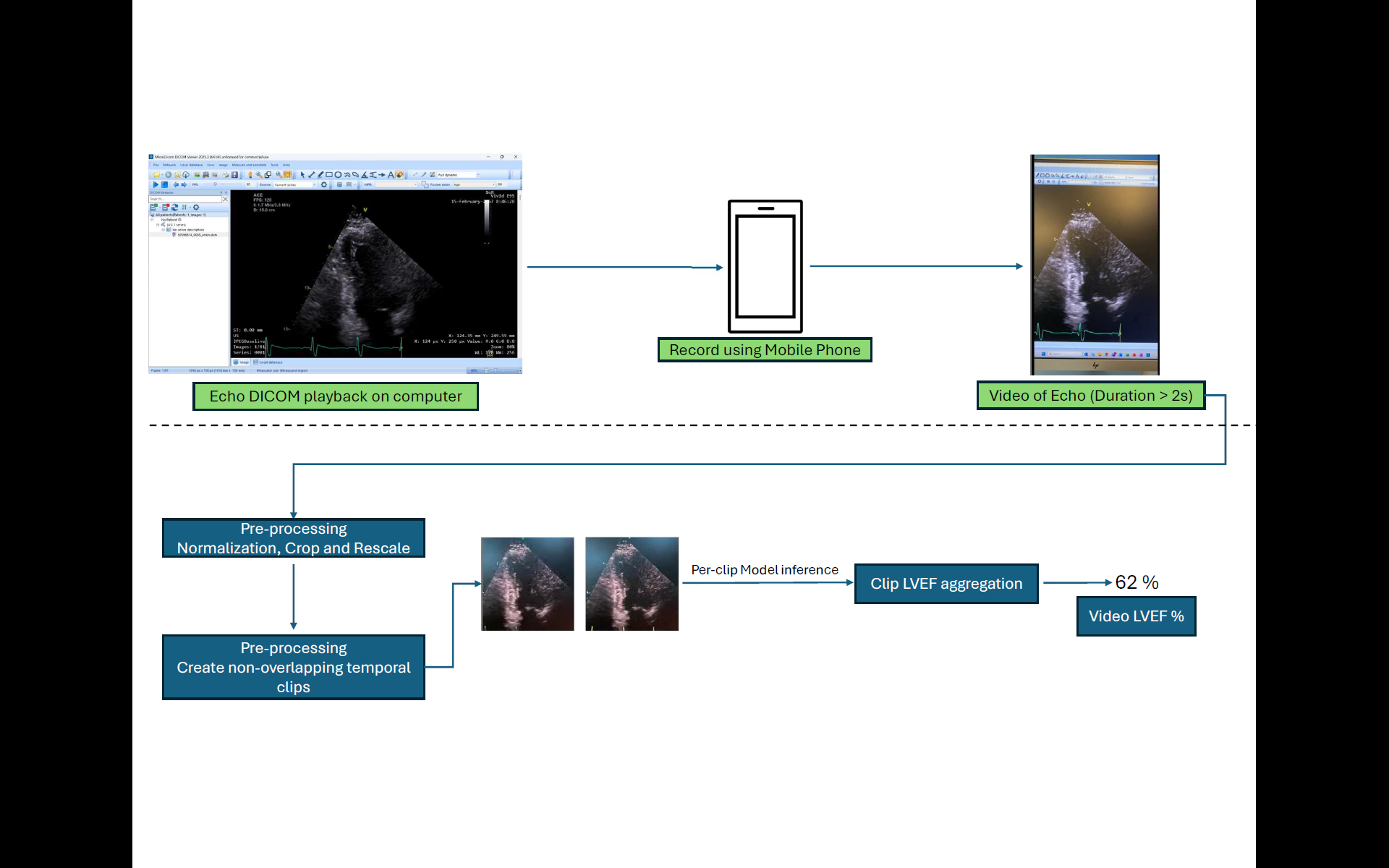

**eFigure 1. Central Illustration Automated Estimation of LVEF on phone recorded videos.**

In this study we evaluated a Pretrained Deep-learning model to Predict LVEF from a dataset of mobile phone recorded videos of echocardiograms. The echocardiograms consisted of A4C, A2C views. The performance of the model was evaluated on 6,209 mobile phone recorded videos consisting of cohorts from a KPNC internal test set, MIMIC-IV-ECHO (BIDMC) and CSMC.

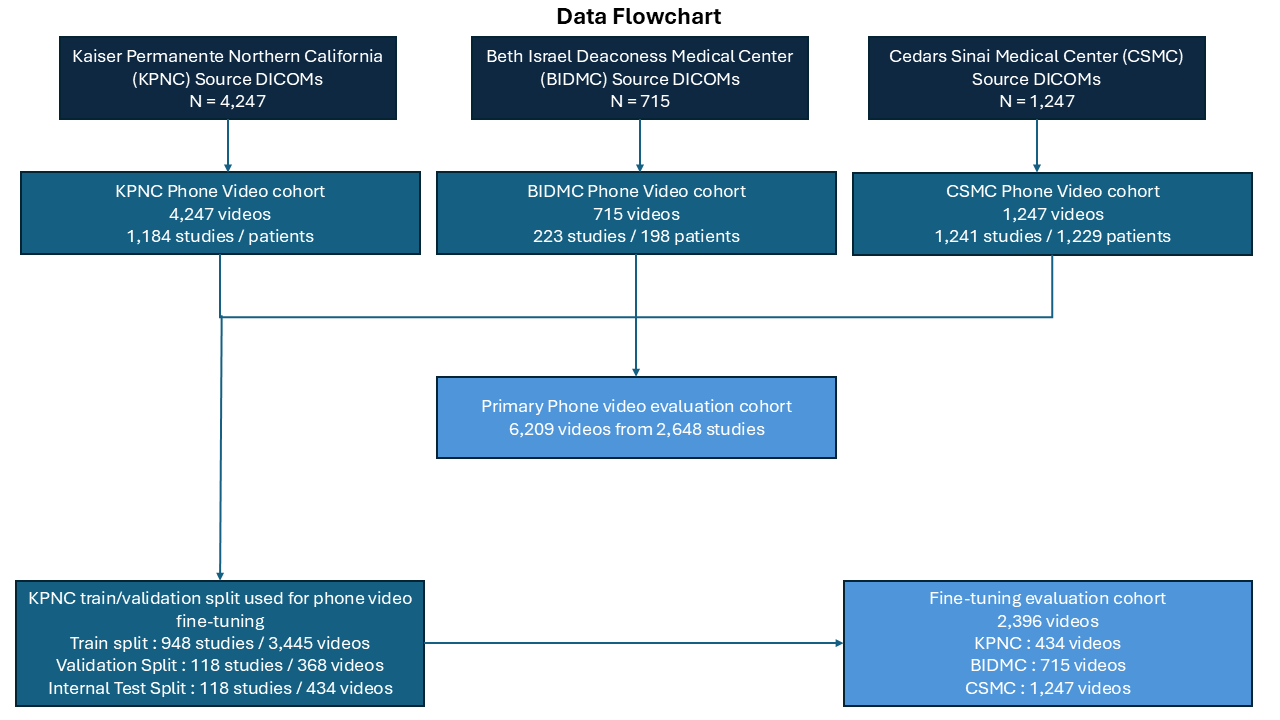

**eFigure 2. Data Flowchart**

Source echocardiographic DICOM videos from Kaiser Permanente Northern California (KPNC), Beth Israel Deaconess Medical Center (BIDMC), and Cedars-Sinai Medical Center (CSMC) were displayed on a computer monitor and recorded using mobile phones to generate site-specific phone-video cohorts. The primary phone-video evaluation cohort included 6,209 videos from 2,648 studies. KPNC videos were split into training, validation, and held-out internal test sets for phone-video fine-tuning. The shared fine-tuning evaluation cohort included the KPNC held-out internal test set and the full BIDMC and CSMC external validation cohorts, for a total of 2,396 videos. Numbers indicate videos unless studies or patients are specified. DICOM, Digital Imaging and Communications in Medicine.

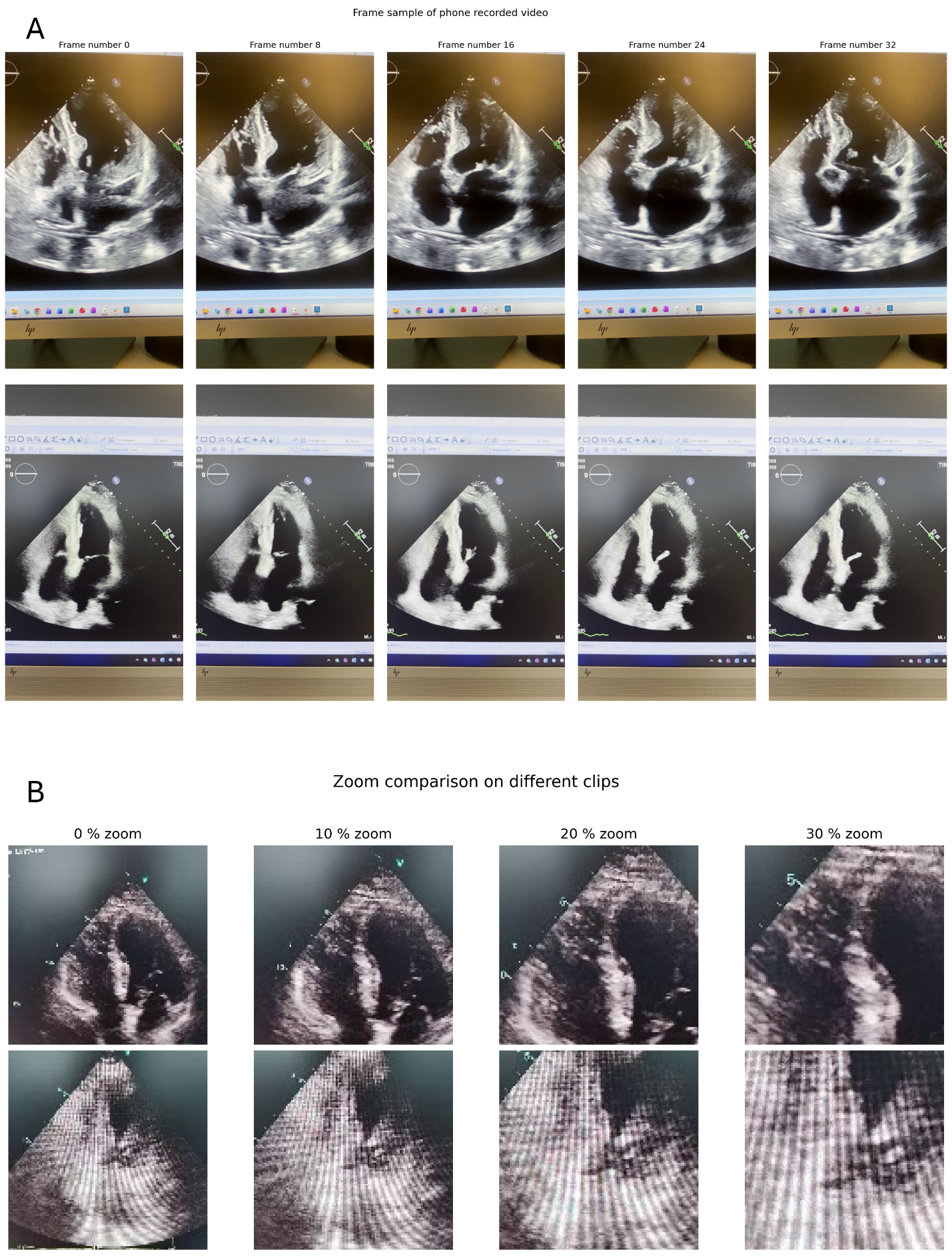

**eFigure 3. Representative mobile-phone-recorded echocardiographic videos and zoom preprocessing conditions.**

(A) Representative frame sequences from multiple mobile-phone-recorded echocardiographic videos at different temporal offsets, illustrating variability in acquisition appearance, including differences in framing, camera angle, glare, display borders and image quality. Despite this variability, key apical cardiac structures remain visible across clips.

(B) Representative examples of progressive zoom preprocessing applied to mobile-phone-recorded echocardiographic videos. Columns show the same clip at 0%, 10%, 20% and 30% zoom. Increasing zoom progressively removes peripheral image context and crops the echocardiographic field, visually corresponding to the observed decline in LVEF prediction performance with tighter framing.
